# Very low levels of physical activity among a broad group of patients hospitalized following hip fracture: A prospective cohort study (the HIP-ME-UP cohort study)

**DOI:** 10.1101/2024.02.09.24302483

**Authors:** Maria Swennergren Hansen, Morten Tange Kristensen, Camilla Kampp Zilmer, Anja Løve Berger, Jeanette Wassar Kirk, Kira Marie Skibdal, Thomas Kallemose, Thomas Bandholm, Mette Merete Pedersen, The HIP-ME-UP Collaborative Group

## Abstract

**Objectives:** The evidence supports early and intensive mobilization and physical activity for hospitalized patients following a hip fracture. Since bedrest and inactivity during acute care are potentially fatal, we need updated knowledge of levels of physical activity in a diverse clinical population. Therefore, the objective was to determine levels of physical activity among a broad representation of patients hospitalized following hip fracture, and secondly to explore the association with 30-day post-discharge readmission, and mortality.

**Design:** Prospective cohort study

**Setting:** Data were collected at two university hospitals in the Capital Region of Denmark from March to June 2023.

**Participants:** Patients hospitalized following hip fracture.

**Main outcome measures:** 24-hr upright time (time standing and walking) was measured from inclusion (post-operative day (POD) 1-3) to discharge using a thigh-worn accelerometer. Readmission and mortality were verified by electronic patient records.

**Results:** 101 patients (62 women) with a mean (SD) age of 79.9 (8.4) years were included. The median (IQR) 24-hr upright time on POD2-6 ranged from 15 (6.9:31.0) to a maximum of 34 (16:67) mins. Patients with cognitive impairment had less upright time than patients without. Post-surgery length of stay was a median of 7 (5:8) days. 25% of the patients were readmitted or had emergency ward referrals and 3% died within 30 days of discharge (no clear association with upright time).

**Conclusions:** Physical activity seems extremely low among a broad representation of patients within the first week following a hip fracture but was not found to influence readmissions. Considering the strong evidence supporting physical activity during acute hospitalization, the low activity level in these patients calls for action.

**Clinicaltrials.gov-identifier**: NCT05756517

## INTRODUCTION

“Physical activity is defined as any bodily movement produced by skeletal muscles that results in energy expenditure” [1]. Clear evidence advocates for physical activity during acute hospitalization, showcasing its benefits on preserving physical functioning, reducing length of stay, and avoiding pulmonary embolism among young and older adults admitted for medical illness [2,3].

A recent systematic review [4] presented low certainty of evidence for a moderate effect of exercise therapy [5] on mobility among patients following a hip fracture, and low evidence for small-to-moderate short-term effects on ADL, lower limb muscle strength and balance [4]. However, for intensified physiotherapy (e.g. 2-3 x daily vs. daily) [6,7] or comprehensive geriatric care [8], the review finds this to be associated with shorter lengths of stay [6], regaining independence in basic mobility [6,7], and increasing upright time during hospitalization [8].

Despite these benefits, physical activity during acute hospitalization, especially in the context of hip fracture, remains challenging [9–11]. Patients with hip fractures often exhibit frailty [12], low muscle mass [13], multiple comorbidities [14], and difficulties in getting out of bed independently [15], making the task even more demanding. Moreover, hospital stays for these patients have become shorter [16], which is why the same tasks must be completed in shorter timeframes.

Research often highlights results from selective populations, excluding individuals with cognitive impairments [17] or language barriers [18]. Considering Denmark’s immigrant population (approximately 13% [19]), a notable portion might not be fluent in Danish. Moreover, around 50% of patients sustaining a hip fracture experience cognitive impairments during hospitalization [20]. Thus, the patient cohorts frequently depicted in the literature fail to represent the reality of clinical settings. Therefore, obtaining results that reflect the true clinical landscape is imperative.

Among patients acutely admitted for medical illness, low physical activity has been linked with an increased risk of 30-day readmission and mortality [21,22]. A similar association was found for patients with a low ambulatory status after a first-time hip fracture [23]. It is uncertain, however, whether increased physical activity levels during hospitalization among patients admitted with hip fractures could yield positive effects on 30-day post-discharge mortality and readmission.

Therefore, the HIP-ME-UP study sought to determine levels of physical activity among a broad representation of patients hospitalized following hip fracture surgery and to explore the association with 30-day post-discharge readmission, and mortality.

## METHODS

### Study Registration and Protocol

HIP-ME-UP was pre-registered on Clinicaltrials.gov (NCT05756517). The present study reports on the first of the two pre-registered objectives. Data on the second pre-registered objective will be published subsequently (data analysis ongoing). We adhered to the STROBE checklist [24] and the REPORT trial guide [25] for reporting of the study.

### Study design and Setting

This prospective cohort study (the HIP-ME-UP cohort) was conducted at two hospitals in the Capital Region of Denmark: Copenhagen University Hospitals Hvidovre (HVH) and Bispebjerg (BBH). We performed the study at two different hospitals to enhance the diversity in patient demographics and hospital settings and to improve external study validity. At HVH, the study was conducted at an orthopedic ward, and at BBH at an ortho-geriatric ward. Data were gathered from March 2023 to June 2023. Data were collected during hospitalization, and patient charts were reviewed 30 days after discharge.

### Patients

We included patients aged 60 years or older, who had undergone surgery for an acute hip fracture at either HVH or BBH, who had a pre-fracture Cumulated Ambulation Score (CAS) (by recall) ≥ 3 points [26], and who demonstrated the ability to provide written informed consent or have a relative or legal guardian available to provide written consent, not later than the third postoperative day (POD). Exclusion criteria: weight-bearing restrictions, multiple fractures or suspected pathological fracture due to cancer, terminal illness or postoperative medical complications limiting the ability to leave the bed, and permanent nursing home residence or unwillingness to cooperate in assessments. For further information please see “Supplementary material”.

#### Treatment

All patients received treatment based on a standardized multimodal fast-track program [27]. Most patients received epidural analgesia pre-operatively [28] until POD4 at HVH and until POD3 at BBH. At HVH, patients received physiotherapy sessions daily from POD1 to POD3, and on most weekdays from POD4. At BBH, patients received physiotherapy on POD1, and continued most weekdays until discharge. The sessions focused on early mobilization, achieving independence in basic mobility (CAS), walking inside with walking aids, stair climbing, and hip-related exercises. Patients were discharged when the treating physician found them medically stable and sufficiently treated for pain. The patients were discharged to their own home or rehabilitation at a municipality-based 24-h care setting based on their level of independence in basic mobility.

#### Patient participation

The Danish version of the written material was read out loud to two patients and their relatives, after which they gave their input, and minor revisions were made. To ensure the inclusivity of patients who did not speak Danish, written material and informed consent were translated by an authorized translation agency into English, Arabic, Urdu, Turkish, and Somali - the five most spoken languages next to Danish at our hospitals. The translated versions were sent to native speaking colleagues at HVH, who read the written material and read it out loud to older relatives. Based on their input, the translated versions were revised to improve understanding. Before formal data collection, we conducted pilot testing on outcome measures involving a sample of five patients.

### Descriptives and Outcomes

An overview of descriptives and outcomes are presented in Table 1. and described below.

**Table. 1.**
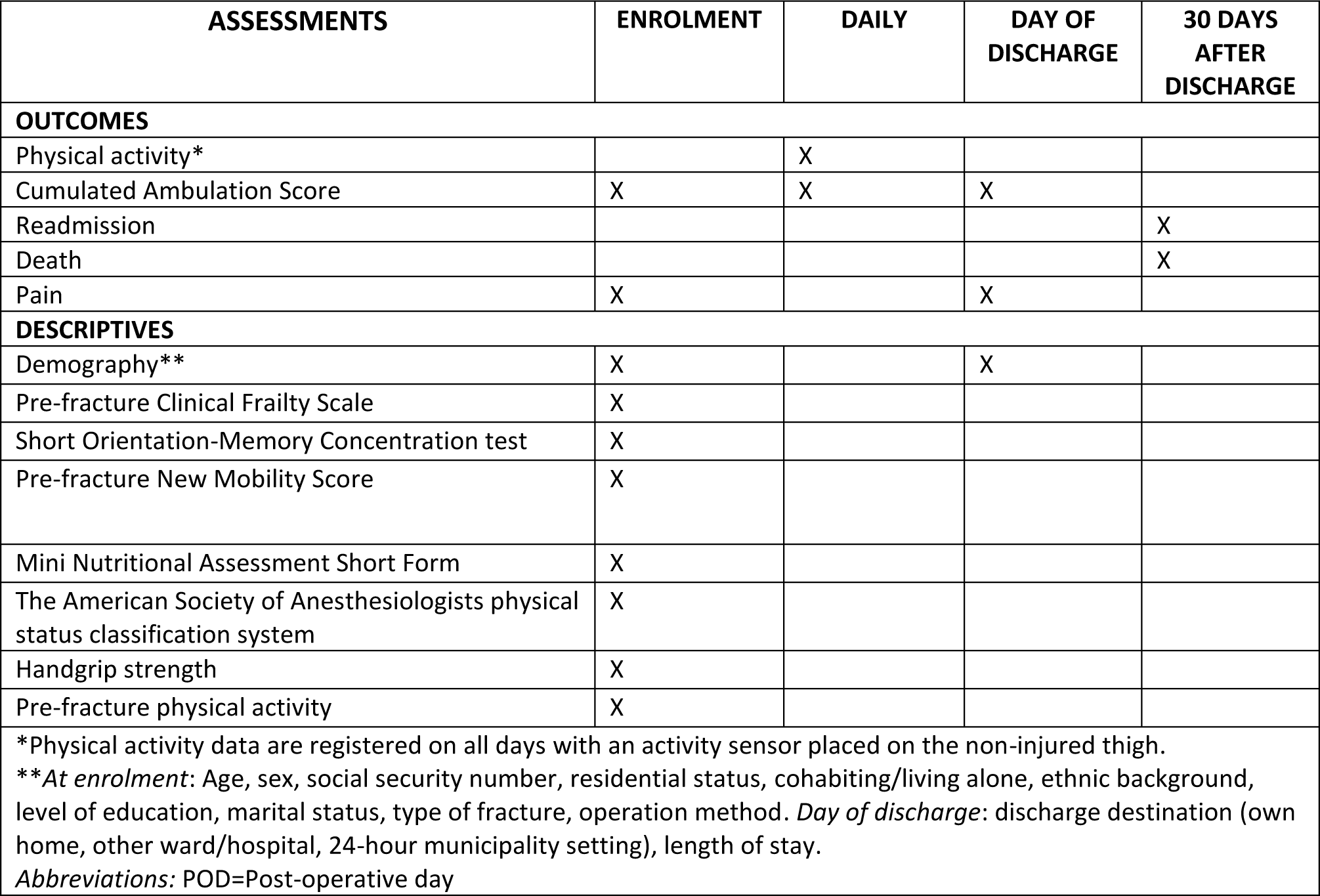
Outcomes, descriptives and timepoints for data collection.

| ASSESSMENTS | ENROLMENT | DAILY | DAY OF DISCHARGE | 30 DAYS AFTER DISCHARGE |
| --- | --- | --- | --- | --- |
| **OUTCOMES** |  |  |  |  |
| Physical activity* |  | X |  |  |
| Cumulated Ambulation Score | X | X | X |  |
| Readmission |  |  |  | X |
| Death |  |  |  | X |
| Pain | X |  | X |  |
| **DESCRIPTIVES** |  |  |  |  |
| Demography** | X |  | X |  |
| Pre-fracture Clinical Frailty Scale | X |  |  |  |
| Short Orientation-Memory Concentration test | X |  |  |  |
| Pre-fracture New Mobility Score | X |  |  |  |
| Mini Nutritional Assessment Short Form | X |  |  |  |
| The American Society of Anesthesiologists physical status classification system | X |  |  |  |
| Handgrip strength | X |  |  |  |
| Pre-fracture physical activity | X |  |  |  |
| *Physical activity data are registered on all days with an activity sensor placed on the non-injured thigh.<br>**At enrolment: Age, sex, social security number, residential status, cohabiting/living alone, ethnic background, level of education, marital status, type of fracture, operation method. Day of discharge: discharge destination (own home, other ward/hospital, 24-hour municipality setting), length of stay.<br>Abbreviations: POD=Post-operative day |  |  |  |  |

#### Primary outcome

*Physical activity* was measured using SENS motion® activity monitors, which are waterproof activity sensors placed laterally on the non-injured thigh (placed in a small patch specially designed for the SENS motion® activity monitor). The SENS motion® monitor records time spent standing and walking (“upright time”), upright events (transitions between sitting and standing), and time spent sitting or lying down (“sedentary time”).

Activity measures were segmented by POD, with calculations performed for each day from 00.00 AM to 11.59 PM. To ensure data completeness, we established a minimum requirement of 23.9 hours of data per POD. For a small number of patients, the SENS motion® monitor was briefly removed for maintenance or adjustment and their data were still included. Further information about the primary outcome is described “Supplementary material”.

#### Secondary outcomes

- *Basic mobility* by the Cumulated Ambulation Score (CAS) [26]. CAS allows day-to-day measurements of basic mobility. It describes the patients’ independence in three activities (getting in and out of bed, sit-to-stand-to sit from a chair with armrests, and indoor walking with or without an assistive device), with each activity assessed on a three-point ordinal scale from 0 to 2 (total 0 to 6 points with 6 indicating independence in basic mobility) [26]. Pre-fracture CAS was based on the patient’s/relative’s self-report on inclusion.
- *Death or readmission 30 days post discharge* assessed via patient journals (any hospital referral was included).
- *Pain* at rest and during walking assessed using the 5-point Verbal Rating Scale (VRS)[29], with categories ranging from “no pain” to “unbearable pain.”

#### Descriptives

Descriptives are listed below and further described in “Supplementary material”.

- *Demographic data* by using questions from the Danish National Health Survey [30]; *Frailty* using the Clinical Frailty Scale (CFS)[31]; *Pre-fracture function* using the New Mobility Score (NMS) [32,33]; *Cognitive impairment* using the Short Orientation-Memory Concentration (OMC) [34,35]; *Nutritional risk* by the Mini Nutritional Assessment Short Form (MNA-SF) [36]; *Health status* using the American Society of Anesthesiologists (ASA) grade [37]; *Body strength* using a test of hand-grip strength (HGS) [38]; and *Pre-fracture physical activity* using a validated questionnaire from the Swedish National Board of Health and Welfare [39].

### Patient inclusion and data collection

Patients were included on weekdays as soon as possible post-operatively and no later than POD3. Two investigators at each hospital included patients and collected data. All with experience with the patient population and with data collection. Information on CAS was collected from the patients’ journals. Registration of CAS is a standard requirement for physiotherapists working at the wards. The staff at the wards were informed about the study before its commencement and were encouraged to ensure continuous monitoring and secure attachment of the activity sensors. The activity sensor was attached immediately at inclusion and removed before discharge - mostly by the investigator and in a few cases by the ward staff. For a more detailed description of the data collection process, visit “Supplementary material”.

### Statistical analysis

We aimed for a sample size of 100 participants, which we considered to be sufficiently representative and in line with samples used in previous studies included in a systematic review of physical activity among older adults after hip fracture [40]. This was a pragmatic decision based on time and resources.

Descriptive data are presented as numbers with percentages, mean with standard deviation, or median with interquartile range, depending on the type and distribution of data. Hourly time spent upright from 7 AM to 11 PM was visually presented by boxplots for POD3-5 to illustrate daily activity patterns concerning patients’ upright positions and walking. We also described the patients on POD4 based on upright time categories (0-15 mins, 15-30 mins. etc.), cognitive impairment, and education.

Logistic regression analysis was used to assess the association between levels of physical activity (upright time, upright events, time walking) on POD4 and risk of 30-day readmission and mortality (in-hospital and 30 days post-discharge), respectively. Both unadjusted, univariable models and adjusted multivariable models were fitted. To adjust for confounding, the analysis included age, sex, fracture type (intracapsular/extracapsular), and CAS (0-5/6) on POD4. For all analyses using physical activity data from POD4, data from POD3 (n=10) were used as surrogate measures if patients had incomplete/missing data on POD4.

As a sensitivity analysis to represent expected extreme cases for the missing data, the regression analysis was repeated using maximum values of complete POD4 data as substitutes for incomplete measures, and the analysis was repeated with minimum values as well. All analyses were performed in R version 4.1.0 [41]. P-values of less than 0.05 were considered statistically significant.

## RESULTS

We screened a total of 267 individuals at HVH and BBH and included 101 patients (Figure 1). Ninety-one patients were able to independently provide consent to participate, while ten patients had a relative or a guardian to provide consent.

**Figure 1.**
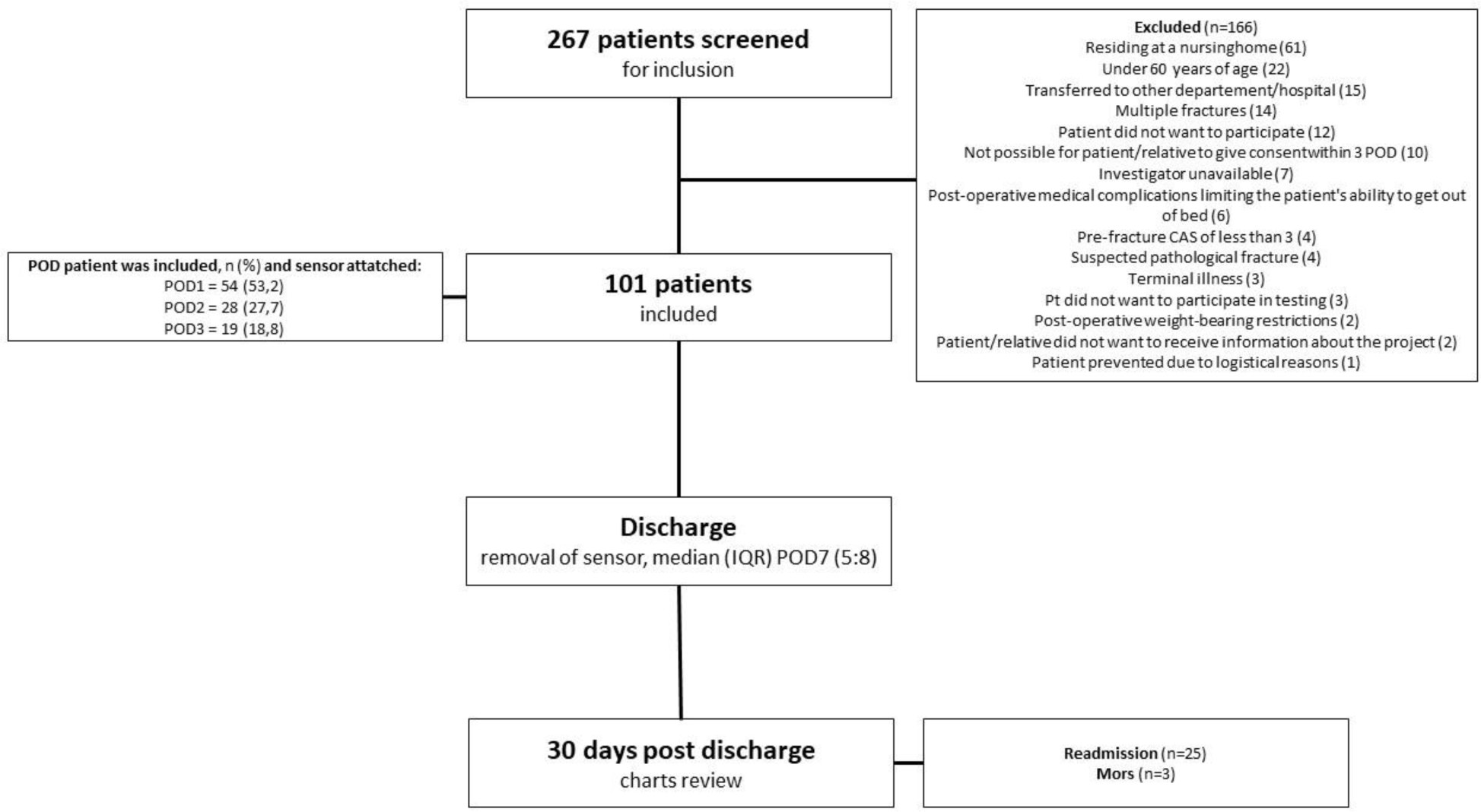
Flowchart of patient inclusion and data collection.

Two activity monitors were lost during the study - one from a patient experiencing delirium, and one that became detached and could not be found. One patient who experienced post-operative medical complications and was destined for palliative care discontinued participation in the study.

Descriptives at enrollment are presented in Table 2, and secondary outcomes at discharge and 30 days post-discharge are presented in Table 3.

**Table 2.** Descriptives at enrollment.

| DESCRIPTIVES AT ENROLLMENT | N=101 |
| --- | --- |
| Age years, mean (SD) | 79.9 (8.4) |
| Women, n (%) | 62 (61.4) |
| Men, n (%) | 39 (38.6) |
| Type of fracture, n (%) |  |
| Intracapsular (cervical femoral fracture) | 54 (53.5) |
| Extracapsular (per- og subtrochanteric) | 47 (46.5) |
| Type of surgery, n (%) |  |
| Parallel pins | 9 (8.9) |
| Hemiarthroplasty | 38 (37.6) |
| Total hip arthroplasty | 2 (2) |
| Dymaic hip screw | 9 (8.9) |
| Short intra medullar hip screw | 31 (30.7) |
| Long intra medullar hip screw | 12 (11.9) |
| **Pre-fracture New Mobility Score** 0-9, median (IQR) | 9 (6:9) |
| **Pre-fracture Cumulated Ambulation Score (CAS)** 0-6, median (IQR) | 6 (6:6) |
| **Pre-fracture independent basic mobility** CAS=6, n (%) | 97 (96%) |
| **Pain at rest** VRS, 1-5, median (IQR) | 1 (1:2) |
| **Pain walking** VRS, 1-5, median (IQR) | 2 (1:3) |
| **The American Society of Anesthesiologists physical status classification system** 1-5, n (%) |  |
| High health status (1-2) | 51 (50.5) |
| Low health status (3-4) | 50 (49.5) |
| **Pre-fracture Clinical Frailty Scale** 1-9, median (IQR) | 3 (3:5) |
| **Short Orientation-Memory Concentration** 0-28, median (IQR) | 20 (15:25,3) |
| **Mini Nutritional Assessment Short form** 0-14, median (IQR) | 8 (6:10) |
| n (%) |  |
| Malnourished (0-7) | 29 (28.7%) |
| At risk of malnutrition (8-11) | 48 (47.5%) |
| Normal nutritional status (12-14) | 0 |
| Missing | 24 (23.7%) |
| **Body Mass Index** median (IQR) | 24 (21:26,2) |
| **Hand grip strength** kg, median (IQR) | 23.6 (17.4:27.2) |
| Women | 20.1 (15.4:24.6) |
| Men | 28.1 (23.6:36.7) |
| **Residential status**, n (%) |  |
| Own home | 99 (98) |
| 24-hour municipality setting | 2 (2) |
| **Cohabiting/living alone**, n (%) |  |
| Cohabiting | 46 (45.5) |
| Living alone | 55 (54.5) |
| **Ethnic background**, n (%) |  |
| Danish | 92 (91.1) |
| From a western country | 6 (5.9) |
| From a non-western country | 3 (3) |
| **Level of education**, n (%) |  |
| Less than 10 years | 35 (34.7) |
| 10-12 years | 19 (18,8) |
| More than 12 years | 47 (46,5) |
| **Marital status**, n (%) |  |
| Married | 44 (43.6) |
| Divorced | 16 (15.8) |
| Widow/widower | 33 (32.7) |
| Unmarried | 8 (7.9) |
| **Pre-fracture level of physical activity 3-19, median (IQR)** | 8 (5:9) |
| **Pre-fracture sedentary time on daily basis, n (%)** |  |
| Virtually all day | 20 (19.8) |
| 13-15 hours | 7 (6.9) |
| 10-12 hours | 18 (17.8) |
| 7-9 hours | 22 (21.8) |
| 4-6 hours | 29 (28.7) |
| 1-3 hours | 3 (3) |
| Never | 0 |
| Missing | 2 (2) |

Abbreviations: IQR = Inter quartile range.

**Table 3.** Secondary outcomes at discharge and 30 days post-discharge.

| OUTCOMES | N=101 |
| --- | --- |
| **Discharge destination, n (%)** |  |
| Own home | 55 (54.5) |
| Another department/hospital | 5 (5) |
| 24-hour municipality setting | 36 (35.6) |
| Homeless shelter | 3 (3) |
| Missing | 2 (2) |
| **Cumulated Ambulation Score (CAS) at discharge 0-6, median (IQR)** | 5 (3:6) |
| **Independent basic mobility at discharge CAS=6, n (%)** | 48 (48.5) |
| **Independent basic mobility (CAS=6) obtained POD, median (IQR)** | 4 (3:6) |
| **POD at which CAS= 6 was registered for those who achieved it, n (%)** |  |
| POD1 | 1 (2.1) |
| POD2 | 7 (14.6) |
| POD3 | 12 (25) |
| POD4 | 6 (12.5) |
| POD5 | 7 (14.6) |
| POD6 | 7 (14.6) |
| POD7 | 4 (8.4) |
| POD9 | 1 (2.1) |
| POD12 | 1 (2.1) |
| **Pain at rest VRS, 1-5, median (IQR)** | 1 (1:2) |
| **Pain at walking VRS, 1-5, median (IQR)** | 3 (2:3) |
| **Length of stay POD, median (IQR) days** | 7 (5:8) |
| **Readmission (any hospital contact), n (%)** | 25 (24.8)<br>1 missing |
| **Reason for readmission, n (%)** |  |
| New fall | 2 (8) |
| Hip related pain | 1 (4) |
| Pulmonary problems | 1 (4) |
| Gastroenterological problems | 1 (4) |
| Suspected hip infection | 5 (20) |
| Hip luxation | 1 (4) |
| Other reasons* | 14* |

\*Others reasons (n): Suspected DVT (5), urinary tract infection (3), large hematoma around the cicatrix (1), accumulation of fluid in the abdominal cavity – not directly correlated to hip surgery (1), low oxygen saturation, increasing shortness of breath and impaired consciousness (1), infarction (1), anemia (1), urinary retention (1). Abbreviations: CAS=Cumulated Ambulation Score; POD=Post-operative day.

### Primary outcome

The median upright time was at most 34 mins (IQR 16:67) on POD6 (n=48), and 24 mins (IQR 11:44) on POD4 (the POD with most included data, n=78). The corresponding median time for walking was 19 mins (IQR 6.5:41) on POD6 and 11 mins (IQR 3.6:28) on POD4. Activity data PODs 2-7 are presented in Table 4 (see end of manuscript). Upright time for more than five minutes per hour only happened between 10-12 am at PODs 3-5. The hourly variation in up-right time is presented in Figure 2.

**Figure 2.**
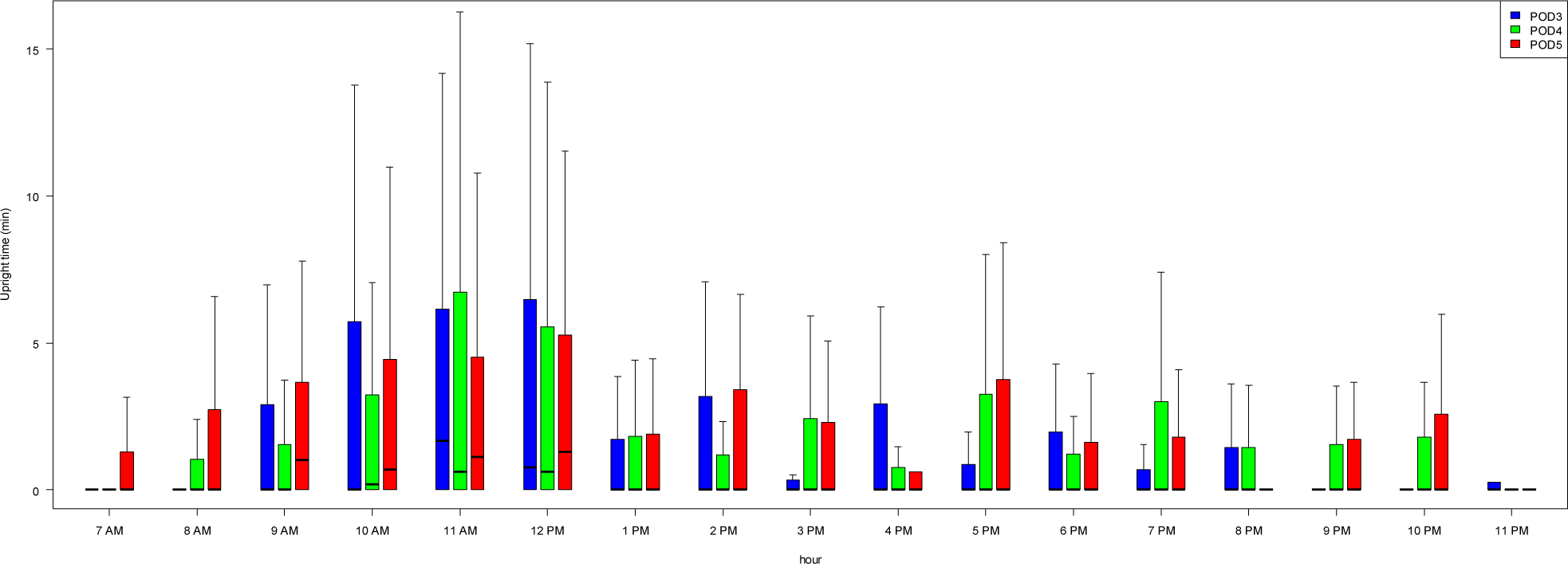
Up-right time (minutes) per hour from 7 am to 11 pm. Blue bar on POD3, green bar on POD4 and red bar on POD5. The thick line in each box indicates median, the colored boxes represent 1. to 3. quantiles, and the thin vertical lines mark extreme data points.

**Table 4.**
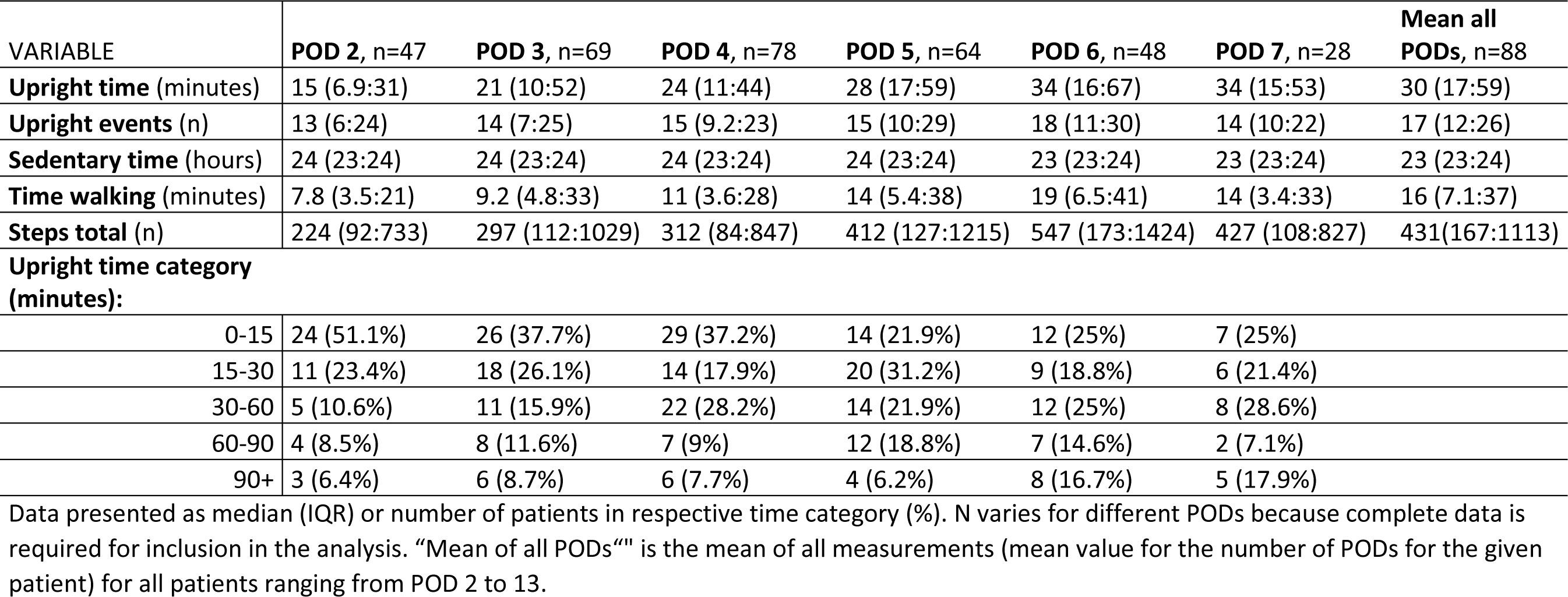
Activity data post-operative days (POD) 2-7.

### Upright time categories

On POD4 (Table 5, see end of manuscript), one-third of the patients (n=31 (35%)) were upright for 15 mins or less. This group of patients was characterized by being malnourished, frail, having low muscle strength, and being dependent on others to get out of bed. Only eight patients (9%) had a median upright time of 90 mins or more. This group of patients was characterized by more being female (88%), not having a cognitive impairment (75%), having age-normative muscle strength [42], having intracapsular fracture (88%), and being independent in basic mobility.

**Table. 5.** Descriptive characteristics within upright time categories on post-operative day (POD) 4, n=89.

| UPRIGHT TIME CATEGORY: | 0-15 minutes<br>n=31 (35%) | 15-30 minutes<br>n=15 (17%) | 30-60 minutes<br>n=23 (26%) | 60-90 minutes<br>n=12 (13%) | 90+ minutes<br>n=8 (9%) |
| --- | --- | --- | --- | --- | --- |
| **DESCRIPTIVE VARIABLE** |  |  |  |  |  |
| **AGE**<br>years, mean (SD) | 80.2 (9.1) | 83.3 (8.2) | 78.3 (8.2) | 80.4 (7.1) | 79 (6.7) |
| **SEX**<br>female, count (% within category) | 20 (65) | 8 (54) | 13 (57) | 7 (58) | 7 (88) |
| **COGNITIVE IMPAIRMENT**<br>yes, count (% within category) | 21 (68) | 11 (73) | 13 (56) | 8 (67) | 2 (25) |
| **HGS FEMALE**<br>kg, median (IQR) | 15.9 (11.2:21.5) | 17.9 (15.0:20.2) | 22.4 (15.4:25.6) | 21.2 (18.6:27.2) | 24.6 (19.1:26.6) |
| **HGS MALE**<br>kg, median (IQR) | 22.8 (17.5:27.3) | 29.5 (23.1:41.4) | 30.8 (23.7:36.3) | 26.0 (25.3:35.2) | 42.5 (42.5:42.5) |
| **PHYSICAL ACTIVITY**<br>pre-fracture 3-19, median (IQR) | 7 (3:8) | 8 (4:9) | 9 (7.75:14.5) | 9 (7.25:12.5) | 10.5 (7.5:16.75) |
| **MNA**<br>0-14, median (IQR) | 7 (6:9) | 8 (6:9.5) | 9.5 (6:10.75) | 10 (8.5:10.5) | 9 (5.75:10) |
| **CFS**<br>1-9, median (IQR) | 5 (3:6) | 4 (3:5) | 3 (3:4) | 3 (3:4.75) | 3 (3.25:3) |
| **FRATURE TYPE**<br>Intracapsular, count (% within category) | 12 (39) | 6 (40) | 11 (48) | 10 (83) | 7 (88) |
| Extracapsular , count (% within category) | 19(61) | 9(60) | 12(52) | 2(17) | 1(12) |
| **ASA**<br>1-2, count (% within category) | 11 (35) | 7(47) | 13 (56) | 7 (58) | 4 (50) |
| 3-4 , count (% within category) | 20 (65) | 8 (53) | 10 (44) | 5 (42) | 4 (50) |
| **CAS POD 4**<br>1-6, median (IQR) | 2 (2:3) | 3 (3:4.5) | 4 (3:6) | 5.5 (3:6) | 6 (5.5:6) |

Data from patients on POD 4 (81 patients, 91%) or POD 3 (8 patients, 8.9%), for those patients with uncomplete data POD4. Pre-fracture physical activity: A score $\geq 11$ corresponds to fulfillment of the World Health Organization's recommendation for weekly physical activity (42). *Abbreviations:* HGS, Handgrip strength; MNA, Mini Nutritional Assessment; CFS, Clinical Frailty Scale; ASA, The American Society of Anesthesiologists; CAS, Cumulated Ambulation Score.

### Patients with cognitive impairments

When looking at physical activity on POD4 and cognition, we found that patients having a cognitive impairment (n= 55) had a median daily upright time of 23.9 mins (IQR 9.9:52.3) and walked a median of 11.3 mins (IQR 3.1:35.0) daily versus 37.8 mins (IQR 14.7:78.3) and 22.7 mins per day (IQR 7.8:51.6), respectively, for patients without a cognitive impairment (n=30).

### Association between activity data POD4 and 30 days readmission and mortality

No clear association was seen between readmission and per minute upright time (OR 1.00, CI 0.98-1.01, p=0.59), per minute time walking (OR 0.99, CI 0.97-1.01, p=0.396), per upright event (OR 1.00, CI 0.96-1.04, p=0.91), or per 100 steps walking (OR 0.98, CI 0.91-1.05, p=0.62). Results from the sensitivity analysis were similar and did not indicate a different interpretation. Because of the low mortality in the sample, the models had trouble converging, resulting in models being unable to produce OR estimates or estimates with extremely wide CI, making estimates useless. Because of this, the result for the mortality analysis is not reported.

## DISCUSSION

In this study, we found very low levels of physical activity on all postoperative days in both an acute orthopedic and an orthogeriatric ward among a broad representation of patients hospitalized following hip fracture surgery. The levels of physical activity were not associated with readmission within 30 days.

According to a systematic review by Zusman et al [40], previous studies investigating physical activity after hip fracture surgery have primarily focused on the period after hospitalization, and many of these with a limited number of patients included [40]. Therefore, the present study adds to the literature by focusing on a broad group of hospitalized patients. The levels of activity found in our study, however, illustrate a possible negative trend in the levels of physical activity among patients hospitalized after hip fracture surgery. In a Norwegian study (2008-2010) on 300 patients Taraldsen et al. [8] compared upright time on POD4 in patients treated in comprehensive geriatric care vs. patients treated in orthopedic care. Their findings favored the comprehensive geriatric care group, showing greater daily upright time (mean 57.6 vs 45.1 mins, p = .016). The average daily upright time was 52 mins (SD 63.7). Similarly, a Danish study (2012) by Kronborg et al. [9] on 37 patients, reported a POD4 median of 42 mins (IQR 9–79). Davenport et al. [10] (2015) found a mean upright time of 16 mins per day among 20 patients, and Haslam-Larmer et al. [11] median upright time of 24 mins (range 0.5-625) among 18 hospitalized patients, whereas we found an upright time of 15 mins (IQR 9.2:23) on POD4. Interestingly, a recent feasibility study in Denmark (2023) comparing physiotherapy twice with once daily (usual care) showed a median upright time of 19.8 mins (IQR 11.4;51.6) on POD4 for the usual care group (n=17) and almost the double for the intervention group (n=29) with a median of 39.6 mins (IQR 15.9;85.5) [7]. Despite the variations in upright time between these studies, and a noticeable negative trend in the levels of physical activity, more intensive physiotherapy can possibly promote physical activity during hospitalization in these patients.

The highest amount of upright time was observed between 10-12 AM on POD3-5 (Fig. 2), with patients maintaining an upright position for approximately 5 minutes per hour. Interestingly, as opposed to our findings, Taraldsen et al [8] found most upright time between 8 AM and 9 AM, when performing morning activities. In our wards, the time slot between 10-12 AM (Fig. 2) often involves physiotherapy sessions for hip fracture patients, and therefore indicates a potential shift from a multidisciplinary approach to a scenario where physiotherapists primarily facilitate patients’ mobility during this limited time, despite established evidence emphasizing the importance of a multidisciplinary treatment approach for these patients [43–45]. This needs to be investigated further. Reflections upon these data have inspired us in our observation strategy in an ongoing ethnographic field study (https://osf.io/3fn59).

Opting not to include patients from nursing homes inadvertently excluded some of the oldest and most comorbid individuals from our study. However, unlike many other studies, our dataset comprised patients with cognitive impairments and those who did not speak Danish as their primary language. This inclusion enabled us to observe that patients with cognitive impairments spend less time upright and less time walking compared to patients without cognitive impairments. This finding aligns with the conclusions drawn in a recent study by Runde et al.[46] that demonstrated that higher cognitive function (overall effect on Mini-Mental State Examination (MMSE)) had a positive impact on upright time the month after surgery. Similarly, in a cohort of acutely hospitalized older patients, patients without cognitive impairment (a MMSE of more than 24) were standing and/or walking significantly more hours during a day than patients scoring less than 24 [47].

Patient treatment during acute hospitalization has evolved, including a shorter length of stay. In our hospital, the day of discharge in 2012 was POD13 (mean ± 6.5) [9]and POD7 (IQR 5:8) in our study. It is noteworthy that both upright time (POD4) and length of stay have halved in our study compared to the study by Kronborg et al. This prompts contemplation about what aspects of treatment are deprioritized in today’s clinical practice.

Increased physical activity during hospitalization has been shown to be associated with reduced 30-day mortality risk among patients without hip fracture hospitalized with pneumonia [22]. However, our study could not explore the link between physical activity and mortality due to considerably lower mortality rates in our samples compared to national levels in Denmark [48].

A previous study highlighted the association between walking activity during hospitalization and 30-day readmission among individuals aged 65 and older hospitalized with acute medical illnesses (OR: 0.90; CI: 0.82-0.98) [21]. We did not observe an association between physical activity and 30-day readmission. A systematic review encompassing predictors of 30-day hospital readmission after hip fracture surgery revealed multifactorial reasons for readmission [49]. Factors like age, ASA grade, individual co-morbidities, and functional status emerged as stronger predictors of readmission risk compared to hospital related factors such as initial length of stay, hospital size, surgical timing, and anesthesia type. Interestingly, medical reasons for readmissions after hip fractures were more common than surgical reasons, similar to what we found in our study’s reported readmissions.

### Limitations

Our dedication to capture a comprehensive patient cohort had constraints. Patients residing in nursing homes were omitted from our study due to their shorter hospital stays. Furthermore, we deemed it ethically inappropriate to involve these individuals and their families since we anticipated potentially limited and often incomplete data from these patients. This exclusion has probably contributed to the relatively low mortality rates observed in our sample, thereby restricting our ability to analyze the relationship between upright time and mortality.

Incorporating patients who did not speak Danish demanded extra resources and effort to translate documents and consent forms. Still, our study only encompassed nine such individuals, limiting our capacity to investigate the potential influence of their non-Danish ethnic backgrounds on their outcomes. Reasons for non-participation among these patients were not documented. Yet, our impression suggests that the few patients declining to participate was not primarily due to language barriers. In fact, we noticed a strong interest and willingness among these patients to contribute, and their relatives provided valuable support.

### Conclusion

Our study revealed concerningly low levels of physical activity among a broad representation of patients hospitalized following hip fracture surgery, which underscores the urgent need for action. The levels of physical activity were not associated with readmission within 30 days. Existing literature emphasizes the advantages of physical activity during hospitalization, not only for patients with hip fracture [2,6,50,51]. We therefore need a deeper understanding and clarification of clinical practices concerning mobility and physical activity, specifically focusing on reasons for lack of activity. This comprehension is crucial to making practical and feasible enhancements in clinical settings.

## DECLARATION OF GENERATIVE AI AND AI-ASSISTED TECHNOLOGIES IN THE WRITING PROCESS

During the preparation of this work MSH used “Open AI/Chat GPT4” to improve readability, spelling, grammar, and the English language in general. After using this tool, all authors reviewed and edited the content as needed and take full responsibility for the content of the publication.

## ETHICAL APPOROVAL

Approval was obtained (Journal-nr.: F-22060655) by the Ethics Committee of the Capital Region of Denmark, and from the Danish Data Protection Agency (P-2023-94). The study adhered to the principles of the Helsinki Declaration.

## Supporting information

Supplementary - coi disclosure

Supplementary - detailed description methods

## Data Availability

The data used in current study might be available (will be assessed individually) from the corresponding author on reasonable request.

## ACKNOWLEDGEMENTS

We are grateful for the support and help from our colleagues, staff, and management in the participating wards at HVH and BBH. Great gratitude to all participating patients and their relatives.

