## Supplementary - detailed description methods for "Very low levels of physical activity among a broad group of patients hospitalized following hip fracture: A prospective cohort study (the HIP-ME-UP cohort study)"

### Supplementary material

#### DETAILED DESCRIPTIONS OF THE METHODS

##### Patients – exclusion criteria

A minor revision of one exclusion criterion "Active cancer or suspected pathological fracture" was made after the inclusion of 18 patients and before any data analysis was conducted to "Suspected pathological fracture due to cancer". This revision was prompted by the discovery of numerous patients living with a current cancer diagnosis, who were otherwise eligible for participation. At the same time, "Terminal illness" was added as an exclusion criterion.

##### About the primary outcome, physical activity:

The sensor is a triaxial accelerometer, which samples at 12 Hz and registers the orientation and acceleration of the thigh. Based on the sensor's orientation and acceleration, a predefined algorithm (version 5.5) categorizes the recordings in predefined activities, e.g., sitting and lying down (X-axis), standing (Y-axis), walking (Z-axis), and number of steps [1].

The SENS motion® system has demonstrated good to high percentage agreement with direct observation for standing, sitting, lying, walking, and the number of steps (all gait speeds). Furthermore, the relative reliability of time spent walking and direct observations was found to be moderate, as indicated by an intraclass correlation coefficient (ICC2.1) of 0.664 (95% confidence interval (CI): -0.003 to 0.903) among older patients with hip fractures [1]. This study [1] identified limitations in detecting slow walking, which for some patients was misclassified as "standing". Consequently, we combined time spent standing and walking into a single variable referred to as "upright time."

**Descriptives:**

- *Demographic data* were collected using questions from the Danish National Health Survey [2]: ethnic background, level of education (<10 years, 10-12 years, >12 years), and marital status (married, divorced, widow, unmarried).
- *Frailty.* Assessed using the Clinical Frailty Scale (CFS), which is a clinical judgment-based frailty scale [3]. The CFS evaluates specific domains including comorbidity, function, and cognition to generate a frailty score ranging from 1 (very fit) to 9 (terminally ill) [3]. The patients were scored by the physiotherapist based on interview of the patient related to their level about 2-weeks preceding their hip fracture [4].
- *Pre-fracture function.* Assessed using the New Mobility Score (NMS) which is a score of a patient's ability to walk indoors, outdoors, and when shopping. It provides a score between 0 and 3 (0: not at all, 1: with help from another person, 2: with a walking aid, 3: no difficulty and no aid) for each

function, and results in a total score from 0 to 9 [5,6]. Being assessed at baseline, the score refers to the week before hospital admission.

- *Cognitive impairment*. Assessed using the Short Orientation-Memory Concentration (OMC). It consists of a 6-item patient reported questionnaire and is validated as a measure of cognitive impairment [7,8]. No distinction was made between patients being cognitively impaired by delirium and those with dementia. A score of 22 or below was considered to reflect cognitive impairment [7].
- *Nutritional risk screening.* Assessed by the Mini Nutritional Assessment Short Form (MNA-SF) [9] which is a validated tool often used within research in this field [10]. It has 6 items and the score ranges from 0 (malnourished) to 14 (normal nutritional status).
- *Health status.* Assessed using The American Society of Anesthesiologists (ASA) grade (35), and categorized into five grades, with 1 indicating a healthy and fit patient and 5 representing a moribund patient not expected to live for 24 hours, with or without surgery.
- *Body strength. A*ssessed using a test of hand-grip strength (HGS). Although the measure of HGS assesses the function of one muscle group, it is regarded as an indicator of overall body strength [12]. HGS was measured using the dominant hand using the highest value of three consecutive tests [13].
- *Pre-fracture physical activity.* Assessed using a validated questionnaire from the Swedish National Board of Health and Welfare, providing a total score from 3 to 19. A score ≥11 corresponds to fulfillment of the World Health Organization's recommendation for weekly physical activity [14].

**Patient inclusion and data collection**

Patients were included on weekdays as soon as possible post-operatively and no later than POD 3. Two investigators at each hospital (MSH, KMS, CKZ, ALB) included patients and conducted the data collection. All with experience with the patient population and with data collection. For the daily registration of CAS, this information was collected from the patients' journal, which is a standardized registration for the physiotherapists working at the wards. The first author (MSH) had the main responsibility for the data collection process. To ensure standardization of the inclusion process as well as assessment of all outcome measures, we reviewed and practiced these procedures among all investigators before the first patients were enrolled. Doubts concerning interpretation of in- and exclusion criteria as well as performance of the outcome measurements were discussed until consensus was reached. The decision on inclusion was based on the investigator's perception of the patient's general condition and the time available for both the investigator and the patient. The staff at the wards were informed about the study before its commencement and were encouraged to ensure continuous monitoring and secure attachment of the activity sensors to mitigate the risk of missing data or sensor loss. After the provision of informed consent, assessments were performed on the wards, and data were entered directly into an electronic database (RedCAP, Research Electronic Data Capture, Vanderbilt University, Nashville, TN, USA) using an iPad. MSH conducted weekly reviews of all typed data to identify any missing data or typing errors. The activity sensor was attached immediately after inclusion by the investigator. The activity sensor was removed before discharge - mostly by the investigator and in a few cases by the ward staff.

It's important to acknowledge that apprehension bias (38), originating from both the staff and the patients, cannot be entirely eliminated in this type of study. In the context of an ongoing research project at the ward, where the primary focus is on physical activity, awareness of this construct is heightened among all involved parties. This heightened awareness may influence how the staff approach and support increased physical activity. To mitigate apprehension bias, efforts were made to encourage the staff to maintain their usual practices and interactions with the patients.
